## Supplementary materials for "Safety, Effectiveness and Immunogenicity of heterologous mRNA-1273 Boost after Prime with Ad26.COV2.S among Healthcare Workers in South Africa: the single-arm, open-label, Phase 3 SHERPA Study"

**Tables**

**Supplementary Table 1: Study schema of Sisonke mRNA-1273 boost and comparator groups**

| **Study Groups** | **Sisonke**  **(Feb–May 2021)** | **Sisonke 2**  **(Nov-Dec 2021)** | **mRNA boost**  **(SHERPA May-Nov 2022)** |
| --- | --- | --- | --- |
| mRNA boost  Group 1 | Ad26.COV2.S | Ad26.COV2.S | mRNA-1273 |
| mRNA boost  Group 2 | Ad26.COV2.S | X | mRNA-1273 |
| Sisonke Comparator Group 1 | Ad26.COV2.S | Ad26.COV2.S | X |
| Sisonke Comparator Group 2 | Ad26.COV2.S | X | X |

**Supplementary Table 2: Eligibility criteria for the SHERPA trial**

**Inclusion criteria**

- Age 18 and older at time of enrolment
- Sisonke participant
- Received a priming Ad26.CoV2.S vaccination as part of the Sisonke study
- If received a second dose of Ad26.CoV2.S vaccine as part of the Sisonke 2 study, this was administered at least 3 months ago
- Participants who are pregnant or report breastfeeding at the time of enrolment may be included.
- Willingness and ability to comply with vaccination plan and other study procedures.
- Capable and willing to provide informed consent

**Exclusion criteria**

- Participants who have received any COVID-19 vaccines other than one or two doses of Ad26.CoV2.S through other means (for example, another mRNA booster dose).
- Current participation in any other research studies (other than Sisonke) that would interfere with the objectives of this study. The determination of whether participation in another study would be exclusionary for a given participant will be made by the PI/designee.
- Occurrence of a known COVID-19 outcome within 14 days of enrollment
- Participants with a history of heparin-induced thrombocytopenia, or thrombosis and thrombocytopenia syndrome.
- History of severe adverse reaction associated with a vaccine and/or severe allergic reaction (e.g., anaphylaxis) to any component of mRNA-1273.
- Pregnancy which began prior to or within 30 days after the receipt of dose 2 of Ad26.COV2 and ongoing at the time of enrolment in this trial.
- Acute infection (e.g. febrile illness) unless resolved before enrolment.
- Any significant acute or chronic medical condition, situation or circumstance that in the opinion of the PI/designee makes the participant unsuitable for participation in the study, or jeopardises the safety or rights of the participant

**The following participants should only be enrolled after discussion with the PSRT**

- Participants who are thought to have suffered a neurological or cardiac adverse event considered related to the Ad26.CoV2.S vaccine.
- Participants reporting a non-infective serious adverse event within the first 28 days following the first or second dose of Ad26.CoV2.S vaccine in the Sisonke trial
- History of myocarditis or pericarditis especially in young male
- Chronic history of severe clotting disorders
- Participants who suffered a thromboembolic adverse event following the Ad26.CoV2.S vaccine.

**Supplementary Table 3: Screening and enrolment visit procedures in the SHERPA study**

| **Heterologous mRNA-1273 Boost Study (Main cohort nested in Sisonke)** | | |
| --- | --- | --- |
| **Visit Number** | **1** | **2** |
| **Study Day** | **-56 to 1** | **1** |
| **Procedure** | **Screen** | **Vaccine** |
| **Study procedures** |  |  |
| Assessment of Understanding | √ |  |
| Informed consent | √ |  |
| Medical history | √ |  |
| Vaccination history | √ |  |
| Physical exam | √ |  |
| Obtain demographics | √ |  |
| Concomitant medications | √ |  |
| Vaccination |  | √ |
| SAEs, AESIs |  | √ |
| **Specimen Collections** |  |  |
| Pregnancy test^#^ | √ |  |
| Blood plasma for SARS CoV-2 serology (4 mls EDTA) |  | √ |
| Nasal swab for SARS CoV-2 PCR |  | √ |

\# Female participants only. Pregnant women were added to the pregnancy registry of the study and had an obstetric and gynaecological history recorded. Follow up was conducted through the sub-study and through the central Sisonke safety desk.

**Supplementary Table 4: Schedule of evaluation for participants in the safety and immunogenicity sub-study (approximately N=200)**

| **Safety and Immunogenicity Sub-study** | | |  |  |
| --- | --- | --- | --- | --- |
| **Visit Number** | **1** | **2** | **3** | **4** |
| **Study Week** |  | 0 | 4 | 24 |
| **Study Day** | -56 to 1 | 1 | 29 | 169 |
| **Procedure** | **Screen** | **Vaccine** |  |  |
| **Study procedures** |  |  |  |  |
| Assessment of Understanding | √ |  |  |  |
| Informed consent | √ |  |  |  |
| Medical history | √ |  |  |  |
| Gynaecological and obstetric history* | (√) |  |  |  |
| Vaccination history | √ |  |  |  |
| Physical exam | √ |  |  |  |
| Obtain demographics | √ |  |  |  |
| Concomitant medications | √ |  |  |  |
| Vaccination |  | √ |  |  |
| SAEs, AESIs assessment |  | √ | √ | √ |
| Early reactogenicity assessment |  | √ |  |  |
| AEs for 28 days post vaccination |  | √ | √ |  |
| **Specimen Collections** |  |  |  |  |
| Pregnancy test ^#^ | √ |  |  |  |
| Blood plasma (approx. 8 mls) |  | √ | √ | √ |
| Blood PBMC (approx. 42 mls)^+^ |  | √ | √ | √ |
| Nasal swab for COVID PCR |  | √ | √ | √ |
| Breastmilk^ for immune responses |  | (√) | (√) | (√) |

^#^female participants only, *pregnant and breastfeeding participants only, ^optional procedure for breastfeeding women, ^+^no peripheral blood mononuclear cells (PBMC) samples were collected from pregnant women

**Supplementary Table 5: Number of SARS-CoV-2 infection and severe Covid-19 events and total exposure time (in years) among SHERPA and non-SHERPA Sisonke participants**

|  | **SHERPA** | | | **non-SHERPA** | | |
| --- | --- | --- | --- | --- | --- | --- |
| **Characteristic** | **Ad26.COV2.S + mRNA-1273** | **2 Ad26.COV2.S + mRNA-1273** | **Total** | **Ad26.COV2.S** | **2 Ad26.COV2.S** | **Total** |
| **SARS-CoV-2 infections** | | | | | | |
| After SHERPA start, but before mRNA-1273 booster | 6 | 13 | 19 | 905 | 2447 | **3352** |
| After mRNA-1273 booster | **2** | **11** | **13** |  |  |  |
| Total | 8 | 24 | 32 |  |  |  |
| Days to infection after mRNA 1273 booster*  Median, IQR | 113.5  (100-127) | 125  (47-164) | 125  (90-154) |  |  |  |
| **Person years at risk#** | | | | | | |
| After SHERPA but before mRNA-1273 booster | 1374.28 | 1682.71 | 3056.99 | 113092.52 | 138012.43 | **251104.95** |
| **After mRNA-1273 booster** | **1553.90** | **1843.96** | **3397.87** |  |  |  |
| **Severe endpoint (COVID-19 hospitalizations or death)** | | | | | | |
| After SHERPA start, but before mRNA-1273 booster | 0 | 0 | 0 | 53 | 95 | **148** |
| After mRNA-1273 booster | 0 | 1 | 1 |  |  |  |
| Total | **0** | **1** | **1** |  |  |  |
| **Person years at risk#** | | | | | | |
| After SHERPA but before mRNA-1273 booster | 1375.42 | 1686.16 | 3061.58 | 113422.48 | 138947.34 | **252369.82** |
| **After mRNA-1273 booster** | **1555.98** | **1848.68** | **3404.66** |  |  |  |

*Events were counted from 14 days after the mRNA-1273 booster. The earliest time an infection occurred in SHERPA was 30 days after vaccination.

#Since the two study groups had dynamic membership and participants could contribute data to both groups, the table presents the percentage of person-years at risk rather than the number of individual participants. SHERPA participants initially included in the non-booster exposure period and left it after receipt of mRNA-1273, provided they did not have confirmed infection.

**Supplementary Table 6: Relative vaccine effectiveness of the mRNA-1273 booster against COVID-19 hospitalizations or death estimates (149 adjudicated endpoints)**

|  | **Unadjusted** | **Adjusted*: Model with comorbidities** | **Adjusted*: Model with HIV** |
| --- | --- | --- | --- |
|  | **VE (95% CI)** | **VE (95% CI)** | **VE (95% CI)** |
| **mRNA-1273 boosted**  **group vs not boosted** | 36%  (-358% to 91%) | 42%  (-319% to 92%) | 38%  (-346% to 91%) |

*Adjusted for age, sex, prior vaccination, prior COVID-19, geographical location

**Supplementary Table 7: Matched cohort analysis matching SHERPA and non-SHERPA participants 1:1 on key variables**

| **Characteristic** | **SHERPA**  **(N=10 503)** | **NON SHERPA**  **(N=10 502)** |
| --- | --- | --- |
| Sex, no.(%) |  |  |
| female | 8445 (80.4) | 8444 (80.4) |
| male | 2058 (19.6) | 2058 (19.6) |
| Median age(IQR), years | 41 (34 - 48) | 41 (34 - 48) |
| Age groups (year), no.(%) | |  |
| 18-39 | 4511 (42.9) | 4511 (42.9) |
| 40-49 | 3815 (36.3) | 3814 (36.3) |
| 50-59 | 1824 (17.4) | 1825 (17.4) |
| 60+ | 353 (3.4) | 353 (3.4) |
| Number of comorbidities, no.(%) | |  |
| 0 | 7332 (69.8) | 7330 (69.8) |
| 1 | 2802 (26.7) | 2803 (26.7) |
| 2+ | 369 (3.5) | 369 (3.5) |
| HIV infection, no.(%) | 1823 (17.4) | 1273 (12.1) |
| Hypertension, no.(%) | 1237 (11.8) | 1592 (15.2) |
| Diabetes mellitus, no.(%) | 437 (4.2) | 557 (5.3) |
| Cancer, no.(%) | 16 (0.2) | 27 (0.3) |
| Tuberculosis, no.(%) | 13 (0.1) | 15 (0.1) |
| Heart disease, no.(%) | 29 (0.3) | 59 (0.6) |
| Chronic lung disease, no.(%) | 15 (0.1) | 43 (0.4) |
| Geographical location, no.(%) |  |  |
| Eastern Cape | 1512 (14.4) | 1513 (14.4) |
| Free State | 446 (4.2) | 446 (4.3) |
| Gauteng | 3544 (33.7) | 3542 (33.7) |
| KwaZulu-Natal | 2611 (24.9) | 2611 (24.9) |
| Limpopo | 138 (1.3) | 138 (1.3) |
| Mpumalanga | 363 (3.5) | 363 (3.5) |
| North West | 337 (3.2) | 337 (3.2) |
| Northern Cape | 4 (0.0) | 4 (0.0) |
| Western Cape | 1548 (14.7) | 1548 (14.7) |

**Supplementary Table 8: Relative Vaccine Effectiveness of the mRNA-1273 booster using the matched cohort analysis approach**

| **SHERPA** | **Non-SHERPA** |  |
| --- | --- | --- |
| **Events/Person Years** | **Events/Person Years** | **Relative Vaccine Effectiveness** |
| 12/3140 | 33/3125 | 63% (32%- 82%) |

**Supplementary Table 9: Multivariable logistic regression model** **of local/systemic reactions adjusted for age and sex**

|  | n (%) reporting AE/reactogenicity | Unadjusted OR  (95% CI) | Adjusted OR  (95% CI) |
| --- | --- | --- | --- |
| **Prior COVID diagnosis** | | | |
| No | 147/ 8372 (1.8%) | Reference |  |
| Yes | 124/ 3424 (3.6%) | 2.10 (1.65 - 2.67) | 2.03 (1.59 - 2.59) |
| **HIV status** | | | |
| No | 234/ 8761 (2.7%) | Reference |  |
| Yes | 37/ 2966 (1.2%) | 0.46 (0.32 - 0.65) | 0.49 (0.34 - 0.69) |
| **Prior vaccination** | | | |
| 1 Ad26.COV2.S | 108/ 5436 (2.0%) | Reference |  |
| 2 Ad26.COV2.S | 163/ 6361 (2.6%) | 1.30 (1.01 - 1.66) | 1.26 (0.99 - 1.62) |

**Supplementary Table 10: Adverse Pregnancy Outcomes among pregnant Women**

| Age range | Time to Onset (days) | Method of Estimation | Outcome | Estimated Gestational Age at Outcome | Delivery Method | Attribution |
| --- | --- | --- | --- | --- | --- | --- |
| 30-34 | 28 | Clinical exam at outcome, LNMP unsure | Ectopic pregnancy | 12 weeks | N/A | Not Related |
| 20-24 | 32 | Date of LNMP | Miscarriage (< 20 weeks) | 12 weeks 1 day | N/A | Not Related |
| 30-34 | 26 | Clinical exam at outcome, LNMP unsure | Miscarriage (< 20 weeks) | 8 weeks 5 days | N/A | Not Related |
| 35-39 | 28 | Date of LNMP | Miscarriage (< 20 weeks) | 14 weeks | N/A | Not Related |
| 44-49 | 6 | Date of LNMP | Miscarriage (< 20 weeks) | 5 weeks | N/A | Not Related |
| 35-39 | 7 | Clinical examination | Miscarriage (< 20 weeks) | 5 weeks | N/A | Not Related |
| 35-39 | 44 | Date of LNMP | Miscarriage (< 20 weeks) | 10 weeks | N/A | Not Related |
| 30-34 | 70 | Date of LNMP | Miscarriage (< 20 weeks) | 14 weeks | N/A | Not Related |
| 30-34 | 56 | Date of LNMP | Miscarriage (< 20 weeks) | 8 weeks | N/A | Not Related |
| 35-39 | 70 | Date of LNMP | Miscarriage (< 20 weeks) | 14 weeks | N/A | Not Related |
| 30-34 | 41 | Clinical examination | Miscarriage (< 20 weeks) | 5 weeks 6 days | N/A | Not Related |
| 30-34 | Not yet pregnant | Date of LNMP | Miscarriage (< 20 weeks) | ≈12 weeks | N/A | Not Related |
| 30-34 | 177 | Date of LNMP | Premature live birth (< 37 weeks) | 31 weeks | C-section | Not Related |
| 25-29 | 19 | Ultrasound | Premature live birth (< 37 weeks) | 29 weeks | C-section | Not Related |
| 35-39 | 82 | Date of LNMP | Premature live birth (< 37 weeks) | 35weeks 5 days | C-section | Not Related |
| 30-34 | 88 | Date of LNMP | Premature live birth (< 37 weeks) | 33 weeks 4 days | C-section | Not Related |
| 35-39 | 154 | Date of LNMP | Spontaneous fetal death and/or still birth (> 20 weeks) | 25 weeks | N/A | Not Related |
| 35-39 | 230 | Date of LNMP | Spontaneous fetal death and/or still birth (> 20 weeks) | 41 weeks | Vaginal | Not Related |
| 30-34 | 41 | Clinical examination | Spontaneous fetal death and/or still birth (> 20 weeks) | 13 weeks 6 days | N/A | Not Related |
| 35-39 | 242 | Date of LNMP | Spontaneous fetal death and/or still birth (> 20 weeks) | 40 weeks | Vaginal | Not Related |
| 35-39 | 112 | Early Ultrasound | Spontaneous fetal death and/or still birth (> 20 weeks) | 31 weeks | Vaginal | Not Related |
| 40-44 | 183 | Date of LNMP | Spontaneous fetal death and/or still birth (> 20 weeks) | 30 weeks | Vaginal | Not Related |
| 30-34 | 70 | Early Ultrasound | Spontaneous fetal death and/or still birth (> 20 weeks) | 38 weeks | C-section | Not Related |

LNMP = Last normal menstrual period; C-section = Caesarean section

**Figures**

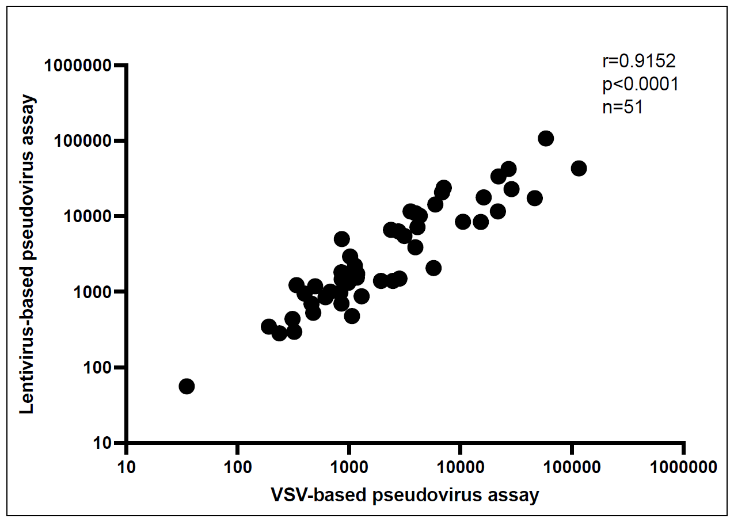

**Supplementary Figure 1: Titer comparison between SARS-CoV-2 lentivirus-based and VSV-based pseudovirus neutralization assays.** Fifty-one samples from HIV-uninfected individuals were tested in both the SARS-CoV-2 lentivirus-based pseudovirus assay and the SARS-CoV-2 VSV-based neutralization assay. Samples were chosen to represent low, middle and high titer values. Titers are depicted as ID50 values for both assays. The correlation between the two assays was measured using the Spearman’s correlation in Graphpad Prism v10.0.2.

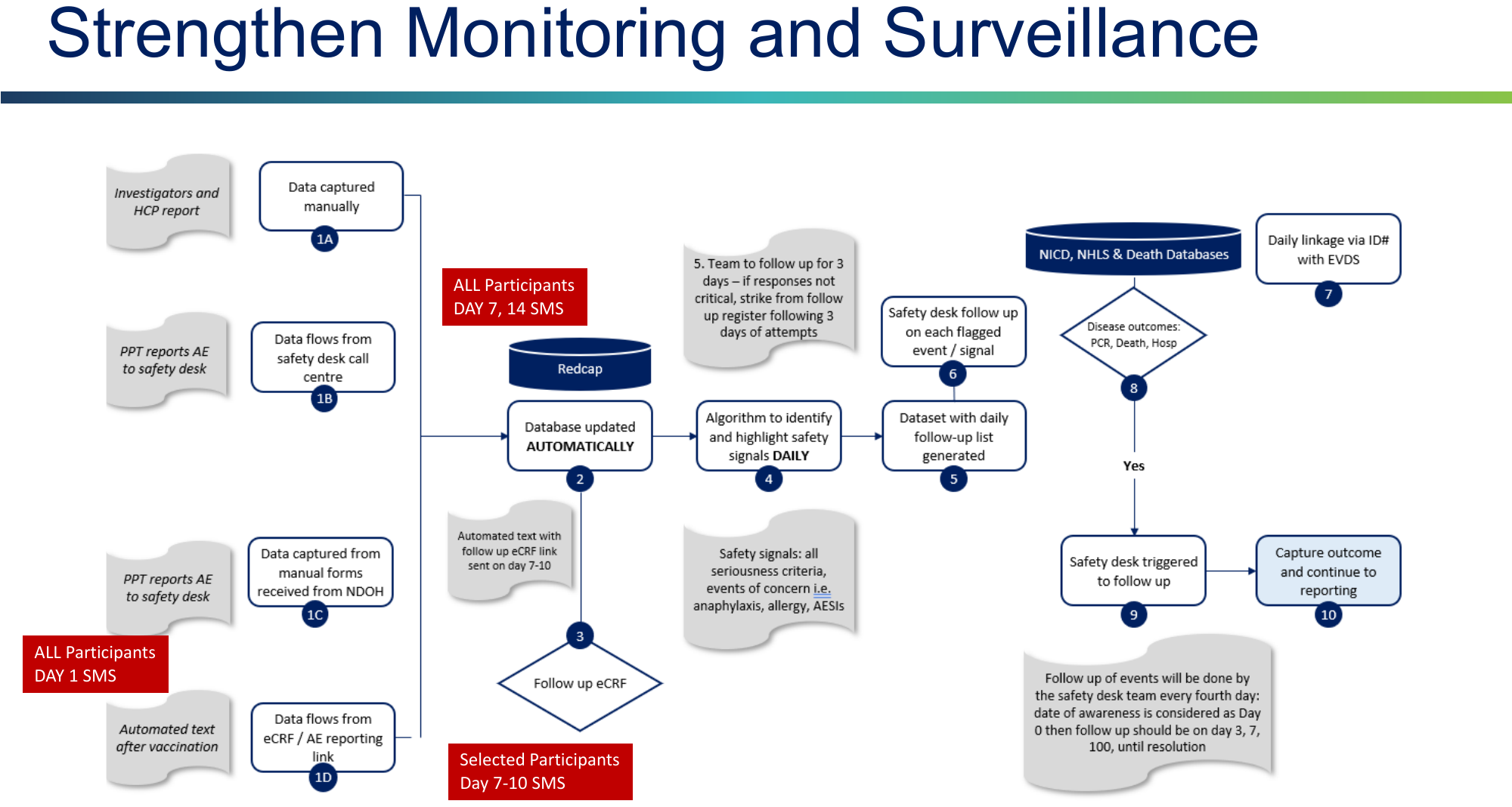

**Supplementary Figure 2: Safety and COVID-19 surveillance in the SHERPA study**

Adjust for confounders

COVID-19 related hospitalizations/ deaths

No COVID-19 related hospitalizations/ alive

No COVID-19 related hospitalizations/ alive

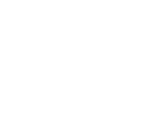

Vaccinated HCWs in Sisonke

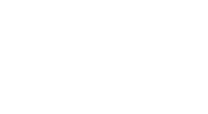

Sisonke Ad26.COV2.S only

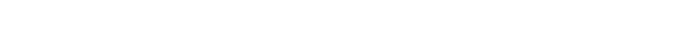

COVID-19 related hospitalizations/ deaths

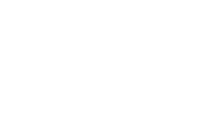

mRNA boosted participants

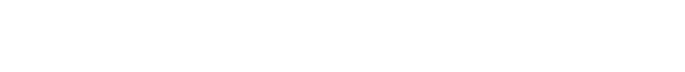

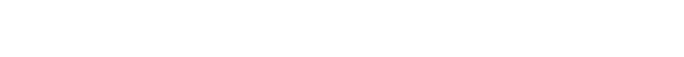

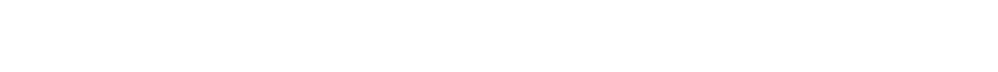

Health care seeking behaviour and selection bias exert an effect. Therefore, **adjusting for potential confounders** is critical

**Supplementary Figure 3: Flow diagram showing the adjusted cohort study design**

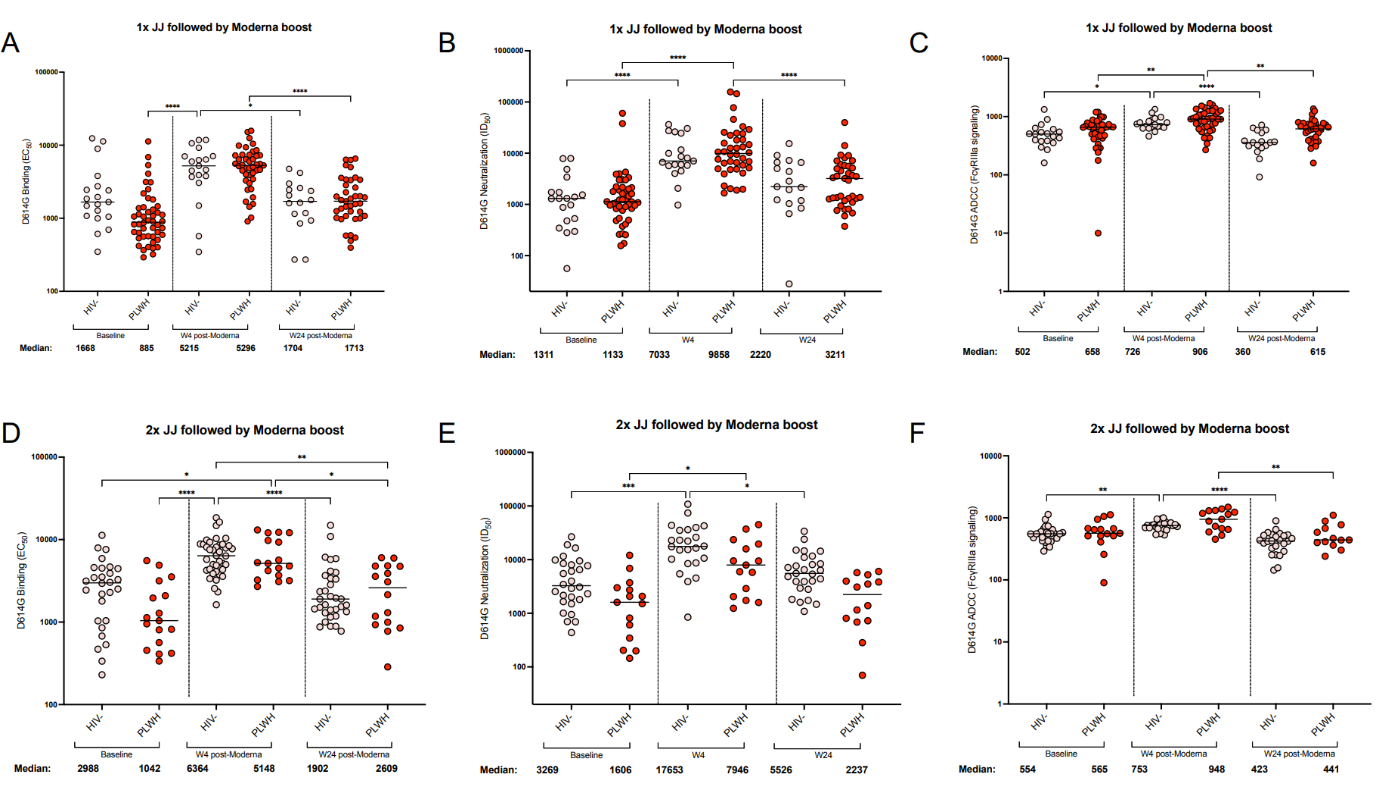

**Supplementary Figure 4: Comparison of antibody response elicited after mRNA-1273 heterologous boost in HIV-negative participants and PLWH.** The binding (Panel A and D), neutralization (Panel B and C) and ADCC (Panel C and F) were measured in individuals who were HIV-negative and people living with HIV (PLWH). Comparisons were separated based on number of Ad26.COV2.S with 1 doses Ad26.COV2.S (Panels A-C) and 2 doses of Ad26.COV2.S (Panels D-F). The plasma neutralization titer is measured as an ID_50_. Black horizontal bars represent medians. The threshold of detection for the neutralization assay is an ID_50_ of 20. Antibody binding was measured using an in-house SARS-CoV-2 assay using the D614G full spike protein. An EC50 was used to measure the binding titers of the samples. ADCC activity was measured by detecting the crosslinking ability of the antibodies present in the serum. Relative light units were measured which correlate with the levels of FcγRIIIa signalling. For all assays, statistical significance was measured with the Kruskal-Wallis test with Dunn’s multiple comparisons test. Significance is shown as: *p < 0.05, **p < 0.01, ***p < 0.001 and ****p < 0.0001. Medians and fold changes are depicted under each graph. Samples were run in duplicate for all assays.

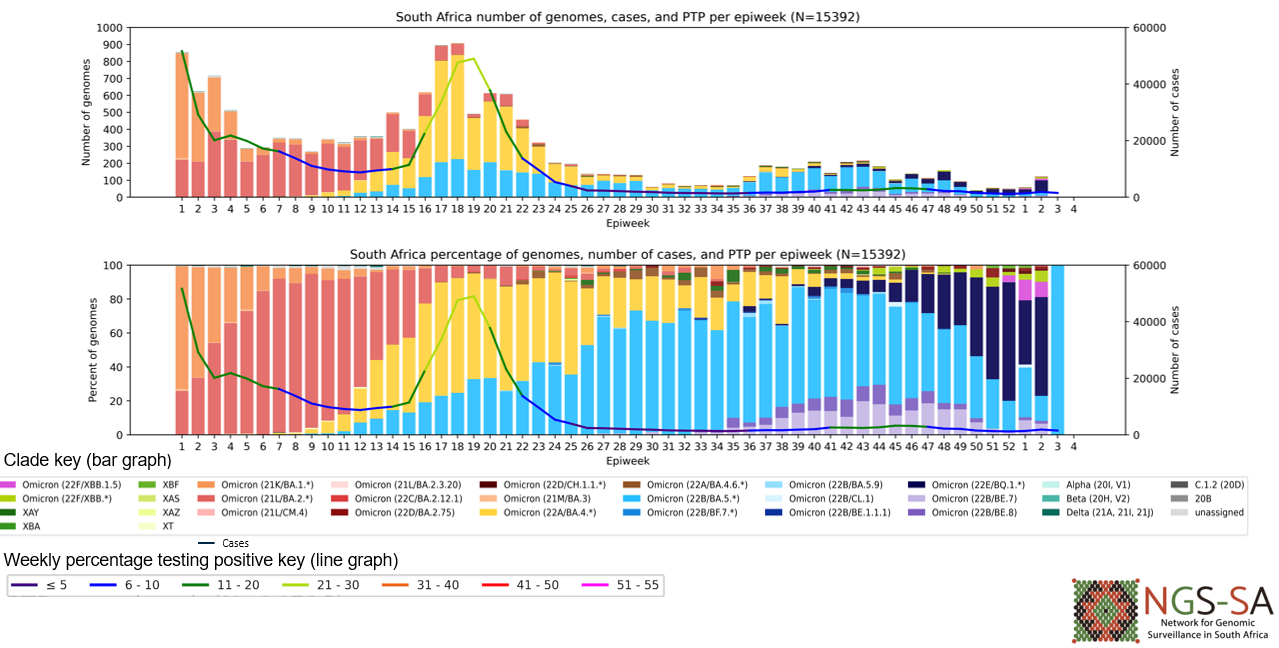

**Supplementary Figure 5: SARS-CoV-2 Infections and circulating Viral strains in South Africa during SHERPA trial 2022-2023 (n=15 392*)**

**Immunogenicity Sub-study Laboratory Assays**

Plasma and PBMCs were collected from a subset of participants at baseline (BL), four weeks (W4) and six months (W24) after the mRNA-1273 booster vaccination. Plasma was used to assess antibody neutralization, binding and antibody dependent cellular cytotoxicity (ADCC). To identify individuals with prior infection or breakthrough infections (BTI) during the study period, we tested all samples for binding against viral nucleocapsid. Of the participants tested, 12/59 and 5/67 of the PLWH were seronegative at BL. When individuals were stratified by Ad26.COV2.S dose, only one participant was seronegative in the one dose Ad26.COV2.S HIV-negative group. Given these small numbers of seronegative participants per group, antibody immunogenicity analyses were only conducted in individuals who were BL seropositive.

*Lentiviral pseudovirus production and neutralization assay*

SARS-CoV-2 pseudotyped lentiviruses were prepared by co-transfecting the HEK 293T cell line with either the SARS-CoV-2 D614G WT (D614G) or Omicron BA.4 (T19I, L24S, Δ25-27, Δ69-70, G142D, V213G, G339D, S371F, S373P, S375F, T376A, D405N, R408S, K417N, N440K, L452R, S477N, T478K, E484A, F486V, Q498R, N501Y, Y505H, D614G, H655Y, N679K, P681H, N764K, D796Y, Q954H, N969K) spike plasmids in conjunction with a firefly luciferase encoding lentivirus HIV-1 pNL4.luc backbone plasmid and incubated at 37 ^o^C for 72 hours. Culture supernatants were clarified using a 0.45-μM filter and stored at −80 °C. For the neutralization assay, the SARS-CoV-2 pseudotyped virus and serially diluted serum samples were incubated for 1 hour at 37°C, 5% CO_2_. Subsequently, 1x10^4^ HEK 293T cells engineered to over-express ACE-2 (293T/ACE2.MF) (kindly provided by M. Farzan (Scripps Research)) were added and incubated at 37°C, 5% CO_2_ for 72 hours upon which luminescence was measured. Titers were calculated as the reciprocal serum dilution (ID_50_) causing 50% reduction of relative light units. Monoclonal antibodies 084-7D, CB6 and CA1 were used as controls.

*Antibody-dependent cellular cytotoxicity (ADCC) assay*

The ability of the serum antibodies to cross-link between CD16 and spike expressed on cells was used as a proxy for antibody-dependent cellular cytotoxicity (ADCC). To express cell surface spike, HEK 293T cells were transfected with SARS-CoV-2 D614G WT-expressing plasmids and incubated at 37°C for 2 days. Spike-expressing cells were then incubated with serum at a 1:100 final dilution in RPMI medium, 10% fetal bovine serum (FBS), and 1% penicillin-streptomycin for 1 hour at 37°C. Jurkat-Lucia NFAT-CD16 cells (Invitrogen) were added to the reaction and incubated for a further 24 hours at 37°C with 10% CO_2_. Signal was read on a luminometer by adding 20 μl of supernatant and 50 μl of QUANTI-Luc secreted luciferase to white 96-well plates. CR3022, P2B-2F6 and Palivizumab served as controls.

*SARS-CoV-2 spike enzyme linked immunosorbent assay (ELISA)*

2 μg/ml of the D614G spike protein was used to coat 96-well, high-binding plates and incubated overnight at 4°C. The plates were incubated in a blocking buffer consisting of 5% skimmed milk powder, 0.05% Tween 20, 1x PBS. Serum samples were diluted to a 1:100 starting dilution followed by a series of 3-fold serial dilutions. Secondary antibody was diluted to 1:3000 in blocking buffer and added to the plates followed by TMB substrate (Thermofisher Scientific). Upon stopping the reaction with 1 M H_2_SO_4_, absorbance was measured at a 450nm wavelength. Monoclonal antibodies CR3022 and palivizumab were used as controls.

*SARS-CoV-2 nucleocapsid enzyme linked immunosorbent assay (ELISA)*

2 ug/ml of nucleocapsid protein (BioTech Africa; Catalogue number: BA25-P) was used to coat 96-well, high-binding plates and incubated overnight at 4 °C. The plates were incubated in a blocking buffer made up of 1x PBS, 5% skimmed milk powder, 0.05% Tween 20. Serum samples were diluted to a 1:100 dilution in blocking buffer and added to the plates as a single dilution. Secondary antibody was diluted to 1:3000 in blocking buffer and added to the plates followed by TMB substrate (Thermofisher Scientific). Upon stopping the reaction with 1 M H_2_SO_4_, absorbance was measured at a 450nm wavelength. Monoclonal antibodies 1A6 and palivizumab were used as controls.

*Measurement of antigen-specific T cells using flow cytometry*

T cell responses to SARS-CoV2 spike were measured as previously described (PMID: 35102311). Briefly, cryopreserved PBMC were thawed, washed and rested in RPMI 1640 (Sigma-Aldrich, St Louis, MO, USA) containing 10% heat-inactivated foetal calf serum for 4 hours prior to stimulation. PBMC were seeded in a 96-well V-bottom plate at ~2 x 10^6^ PBMC per well and stimulated with a commercial ancestral SARS-CoV-2 spike (S) pool (1 µg/mL, Miltenyi Biotec, Surrey, UK) or variant spike mega pools (15 mers with 10-aa overlap) spanning the entire S of the ancestral, Omicron BA.1 and XBB.1 variants (1 µg/mL). All stimulations were performed in the presence of Brefeldin A (10 µg/mL, Sigma-Aldrich) and co-stimulatory antibodies against CD28 (clone 28.2) and CD49d (clone L25) (1 µg/mL each; BD Biosciences, San Jose, CA, USA). As a negative control, PBMC were incubated with co-stimulatory antibodies, Brefeldin A and an equimolar amount of DMSO. After 16 hours of stimulation, cells were washed, stained with LIVE/DEAD™ Fixable Near-IR Stain (Invitrogen, Carlsbad, CA, USA) and subsequently surface stained with the following antibodies: CD14 APC-Cy7 (HCD14, Biolegend, San Diego, CA, USA), CD19 APC-Cy7 (HIB19, Biolegend), CD4 BV785 (OKT4, Biolegend), CD8 FITC, CD45RA BV570 (HI100, Biolegend), CD27 PE-Cy5 (1A4, Beckman Coulter, Brea, CA, USA). Cells were then fixed and permeabilized using a Cytofix/Cyto perm buffer (BD Biosciences) and stained with CD3 BV650 (OKT3), IFN-γ BV711 (4S.B3), TNF-α PE-Cy7 (Mab11) and IL-2 PE/Dazzle^™^ 594 (MQ1-17H12) from Biolegend. Finally, cells were washed and fixed in CellFIX (BD Biosciences). Samples were acquired on a BD Fortessa flow cytometer and analyzed using FlowJo (v10.8.1 FlowJo LLC, Ashland, OR, USA). Results are expressed as the frequency of total memory CD4 or CD8 T cells expressing IFN-γ, TNF-α or IL-2. Due to high TNF-α backgrounds, cells producing TNF-α alone were excluded from the analysis. All data are presented after background subtraction.
